# Prevalence and real-world effectiveness of popular smoking cessation aids in England: a population-based study

**DOI:** 10.1101/2024.09.16.24313731

**Authors:** Sarah E. Jackson, Jamie Brown, Vera Buss, Lion Shahab

**Affiliations:** Department of Behavioural Science and Health, University College London, London, UK; SPECTRUM Consortium, UK

**Keywords:** smoking cessation, quitting, quit success, treatment, observational

## Abstract

**Importance:** A wide range of medications, non-combustible nicotine products, behavioural support, and alternative treatments are available in England to help people stop smoking. Understanding their effectiveness in the real world can support informed decision-making.

**Objectives:** To provide up-to-date estimates of the prevalence and real-world effectiveness of different smoking cessation aids and explore moderation of effectiveness by socioeconomic position.

**Design:** Population-based survey, 2006-2024.

**Setting:** England.

**Participants:** 25,094 adults (≥16y) who reported having tried to quit smoking in the past year.

**Main outcomes and measures:** The outcome variable was self-reported continuous abstinence from the start of the most recent quit attempt up to the time of survey. Independent variables were use (yes/no) of the following aids in the most recent attempt: nicotine replacement therapy (NRT) obtained on prescription or over-the-counter; varenicline; bupropion; e-cigarettes; face-to-face behavioural support; telephone support; written self-help materials; websites; smartphone apps; hypnotherapy; Allen Carr’s Easyway method; heated tobacco products (HTPs); nicotine pouches. Covariates included sociodemographic characteristics and features of the quit attempt.

**Results:** We analysed data from 25,094 participants (mean [SD] age = 38.7y [15.3]; 48.5% women). In 2023/24, the most used aids were e-cigarettes (40.2%) and over-the-counter NRT (17.3%); 40.8% of quit attempts were unaided. While e-cigarette use was associated with higher odds of abstinence after adjustment for use of other aids and covariates (OR=1.95 [1.74-2.17]), use of over-the-counter NRT was not (OR=1.03 [0.93-1.15]). Other aids positively associated with abstinence were websites (used by 4.6% in 2023/24; OR=1.43 [1.03-1.98]), prescription NRT (4.5%; OR=1.33 [1.12-1.58]), varenicline (1.1%; OR=1.80 [1.50-2.18]), and HTPs (0.7%; OR=2.37 [1.24-4.51]). Face-to-face behavioural support (used by 2.2% in 2023/24) was also associated with higher odds of abstinence among those from less advantaged (OR=1.59 [1.19-2.14]) but not more advantaged social grades (OR=0.91 [0.65-1.29]). There was not clear evidence of a benefit of any other aid, although some analyses were inconclusive.

**Conclusions and relevance:** A range of effective smoking cessation aids are available in England, but many people try to quit using less effective forms of support or none at all. Quit success rates could be improved by encouraging people to use more effective methods.

## Introduction

Stopping smoking is one of the best things a person can do to improve their health, but can be very difficult.^1^ Nicotine delivered via cigarettes and other combustible tobacco products is highly addictive and despite people’s best intentions, most quit attempts fail.^2^ Failure rates are typically higher among people from more disadvantaged socioeconomic groups,^3^ contributing to health inequalities.

A wide range of aids are available to help people stop smoking. These include medications (e.g., nicotine receptor partial agonists), non-combustible nicotine products (e.g., nicotine replacement therapy [NRT], e-cigarettes), behavioural support (e.g., face-to-face, digital), and alternative treatments (e.g., hypnotherapy). Many of these have been found to increase quit success rates in randomised controlled trials (RCTs).^4–8^ However, trial results do not always replicate in real-world settings, so findings must be triangulated with observational evidence.^9^

In England, the best estimates of the real-world effectiveness of different smoking cessation aids are derived from the Smoking Toolkit Study. This is a nationally-representative monthly cross-sectional survey that has been collecting detailed data on smoking and smoking cessation since 2006.^10^ In a series of papers, we have analysed data accumulated from a growing sample of smokers to compare the success rates of those trying to quit with different aids, adjusting for a wide range of potential confounders (e.g., level of addiction and other features of the quit attempt).^11–16^ The largest analysis to date included 18,929 participants surveyed up to July 2018 and suggested using e-cigarettes or varenicline were the most effective methods: after adjustment for covariates and use of other aids, participants who used these in their most recent past-year quit attempt had 1.95 and 1.82 times higher odds, respectively, of still being abstinent at the time of the survey than those who did not.^11^ There was little evidence that effectiveness differed by socioeconomic position, with the exception of websites, which appeared more effective for smokers from more disadvantaged social grades compared with those from more advantaged social grades.^11^ However, results were insensitive for most of the other aids studied, meaning more data are required to draw firmer conclusions on their real-world effectiveness.^11^

This study aimed to provide a comprehensive update on the real-world effectiveness of smoking cessation aids in England. Using data collected between 2006 and 2024, we updated our previous estimates for prescription NRT, NRT bought over the counter, varenicline, bupropion, e-cigarettes, face-to-face behavioural support, telephone support, written self-help materials, websites, and hypnotherapy. In addition, we extended the analysis to cover other types of behavioural support (smartphone apps and Allen Carr’s Easyway method [a pharmacotherapy-free behavioural programme]) and newer non-combustible nicotine products (heated tobacco products and nicotine pouches). We also tested for moderation of treatment effectiveness by socioeconomic position and provided data on the prevalence of use of each cessation aid across the study period.

## Methods

### Pre-registration

The study protocol and analysis plan were pre-registered on Open Science Framework (https://osf.io/uyw5k/). We also ran some unplanned analyses on the prevalence of smoking cessation aid use to provide contextual information (see *statistical analysis* section for details).

### Design

The Smoking Toolkit Study is an ongoing monthly cross-sectional household survey.^10^ It uses a hybrid of random probability and simple quota sampling to select a new representative sample of approximately 1,700 adults (≥16 years) in England each month.^10^ Data have been collected face-to-face up to the start of the Covid-19 pandemic and via telephone since April 2020.^17^ Data were not collected from 16- and 17-year-olds between April 2020 and December 2021.

We analysed data collected in the period from November 2006 (the first wave of data collected) to June 2024 (the most recent data at the time of analysis).

### Sample selection

We selected participants who reported: (i) smoking cigarettes or any other combustible tobacco product daily or occasionally at the time of the survey or during the preceding 12 months (‘past-year smokers’) and (ii) having made at least one serious quit attempt in the preceding 12 months.

### Measures

Full details of the measures are provided in the Supplementary File.

### Outcome: quit success

The outcome variable was self-reported continuous abstinence from the start of the most recent quit attempt up to the time of survey.

### Exposures: use of cessation aids

Independent variables were self-reported use or not (dummy coded) of the following smoking cessation aids in the most recent quit attempt: prescription NRT; NRT bought over the counter; varenicline (Champix); bupropion (Zyban); e-cigarettes; face-to-face behavioural support; telephone support; written self-help materials; websites; smartphone apps; hypnotherapy; Allen Carr’s Easyway method (delivered face-to-face); heated tobacco products; nicotine pouches. Participants were asked to indicate all that apply, and data for each were coded 1 if chosen and 0 otherwise.

Some aids were not assessed in every wave: written self-help materials were included in the list of options from March 2007, websites and Allen Carr’s Easyway from April 2008, e-cigarettes from July 2009, smartphone apps from February 2012, heated tobacco products from April 2016, and nicotine pouches from June 2021. We imputed missing values as 0 for participants surveyed before the response options were introduced (because each product was very rarely used immediately after each measure was added).

### Covariates

Covariates included age, gender, occupational social grade (more advantaged ABC1 vs. less advantaged C2DE^18^), level of addiction,^19^ time since the quit attempt started, the number of prior quit attempts in the past year, whether or not the quit attempt was planned, whether the participant cut down first or stopped abruptly, calendar month, survey year, and mode of data collection.

### Statistical analysis

We analysed the data using R v.4.2.1. Survey weights were applied so that the sample matched the demographic profile of adults in England.^10^ All analyses were done on complete cases.

### Prevalence of use (unplanned)

We reported the proportion of participants who reported using each of the aids. Because our study spanned a 17.5-year period, during which the availability and use of the various aids differed, we graphically displayed the proportions using each aid across the study period (calculated as 3-month moving averages, to reduce noise) to provide context on changes over time. To provide up-to-date estimates of aid use, we separately reported the proportion of participants surveyed in 2023/24 who reported using each aid.

### Associations with abstinence (pre-registered)

We calculated the quit success rate (with 95% confidence interval [CI]) among users of each aid. Then we used logistic regression to analyse associations between quit success (abstinent yes vs. no) and use of different smoking cessation aids (use vs. no use of a specific aid). We ran three models. Model 1 included all other cessation aids (to estimate the unique association between each cessation aid and abstinence), but no covariates. Model 2 included covariates, but no other cessation aids. Model 3 was fully adjusted for all cessation aids plus covariates.

To test for moderation of effectiveness by socioeconomic position, we repeated Model 3 in a series of fully adjusted models in which two-way interactions between the cessation aids and occupational social grade (ABC1 vs. C2DE) were added. The interaction with occupational social grade was tested in a separate model for each cessation aid. Where there was evidence of moderation of treatment effectiveness, we reran Models 1-3 in stratified analyses to provide more information as to the nature of the differences between groups.

As we have done previously,^11^ we calculated planned Bayes factors (using an online calculator^20^) for non-significant associations between use of a given aid and quit success, to determine whether the data provided evidence that the null hypothesis was more likely than a specified effect or were insensitive to distinguish between the two.^21,22^ The specified effect was represented by a half-normal distribution reflecting an effect of OR=1.5 (a conservative estimate in the ballpark of interventions known to be effective^23^; determined *a priori* and pre-registered). Values□≥3 can be interpreted as evidence that the effect is more likely than the null, □≤1/3 as evidence the null is more likely, and between 1/3 and 3 suggest the data are insensitive to distinguish between _them._21,22

## Results

Data were collected from 77,372 past-year smokers, of whom 26,789 (34.6%) reported having tried to quit in the past year. We excluded 1,695 (6.3%) with missing data on one or more variables, leaving a final sample of 25,094 participants (weighted mean [SD] age = 38.7 [15.3] years; 48.5% women). There were more missing data since the interviews switched from face-to-face to telephone, but the analysed sample was broadly similar to those who were excluded in their sociodemographic characteristics and the features of their quit attempt (**Table S1**).

### Prevalence of use

Just over half (55.8%) of participants reported using at least one cessation aid in their most recent quit attempt. Across the period, the most commonly used aids were over-the-counter NRT (24.5%) and e-cigarettes (19.0%), followed by prescription NRT (7.0%), varenicline (4.8%), and face-to-face behavioural support (3.8%; **Table 1**).

**Table 1.** Prevalence of use of cessation aids in England, overall and in 2023/24.

| Cessation aid used in most recent quit attempt<sup>1</sup> | Overall |  | 2023/24<sup>3</sup> |
| --- | --- | --- | --- |
|  | N<sup>2</sup> | % [95% CI] | % [95% CI] |
| E-cigarettes<sup>4</sup> | 4,459 | 19.0 [18.4–19.5] | 40.2 [37.6–42.8] |
| Over-the-counter NRT | 6,258 | 24.5 [23.9–25.1] | 17.3 [15.3–19.2] |
| Websites<sup>4</sup> | 430 | 1.9 [1.7–2.0] | 4.6 [3.5–5.7] |
| Prescription NRT | 1,842 | 7.0 [6.6–7.3] | 4.5 [3.4–5.5] |
| Smartphone apps<sup>4</sup> | 186 | 0.8 [0.7–0.9] | 3.6 [2.6–4.7] |
| Nicotine pouches<sup>4</sup> | 84 | 0.4 [0.3–0.5] | 3.1 [2.2–4.1] |
| Written self-help materials<sup>4</sup> | 425 | 1.8 [1.6–2.0] | 2.3 [1.5–3.0] |
| Face-to-face behavioural support | 1,013 | 3.8 [3.6–4.1] | 2.2 [1.5–2.9] |
| Telephone support | 224 | 0.9 [0.8–1.0] | 1.3 [0.8–1.9] |
| Hypnotherapy | 213 | 0.9 [0.7–1.0] | 1.2 [0.7–1.8] |
| Varenicline | 1,208 | 4.8 [4.5–5.1] | 1.1 [0.5–1.7] |
| Bupropion | 346 | 1.3 [1.2–1.5] | 0.9 [0.4–1.5] |
| Heated tobacco products<sup>4</sup> | 72 | 0.3 [0.2–0.3] | 0.7 [0.3–1.1] |
| Allen Carr's Easyway<sup>4</sup> | 45 | 0.2 [0.1–0.3] | 0.5 [0.1–0.9] |
| None of these | 11,079 | 44.2 [43.5–44.9] | 40.8 [38.2–43.4] |

CI, confidence interval.
<sup>1</sup> Not mutually exclusive; sorted from the most to least popular in 2023/24.
<sup>2</sup> Unweighted number of participants who reported using each aid.
<sup>3</sup> Up-to-date estimates of prevalence of the use of each aid among participants surveyed between January 2023 and June 2024 (*n*=1,642).
<sup>4</sup> Use of written self-help materials was assessed from March 2007, websites and Allen Carr's Easyway from April 2008, e-cigarettes from July 2009, smartphone apps from February 2012, heated tobacco products from April 2016, and nicotine pouches from June 2021; use was imputed as 0 before this.

However, patterns of aid use changed over time (**Figure 1**; **Figure S1**). There was a large increase in the use of e-cigarettes, which became the most popular aid used in 2013. There were also smaller increases in the use of novel non-combustible nicotine products (in particular, nicotine pouches) and digital support (websites and smartphone apps) in more recent years. There were decreases in the use of over-the-counter NRT, prescription medications (varenicline, NRT, and bupropion), and face-to-face behavioural support.

**Figure 1.**
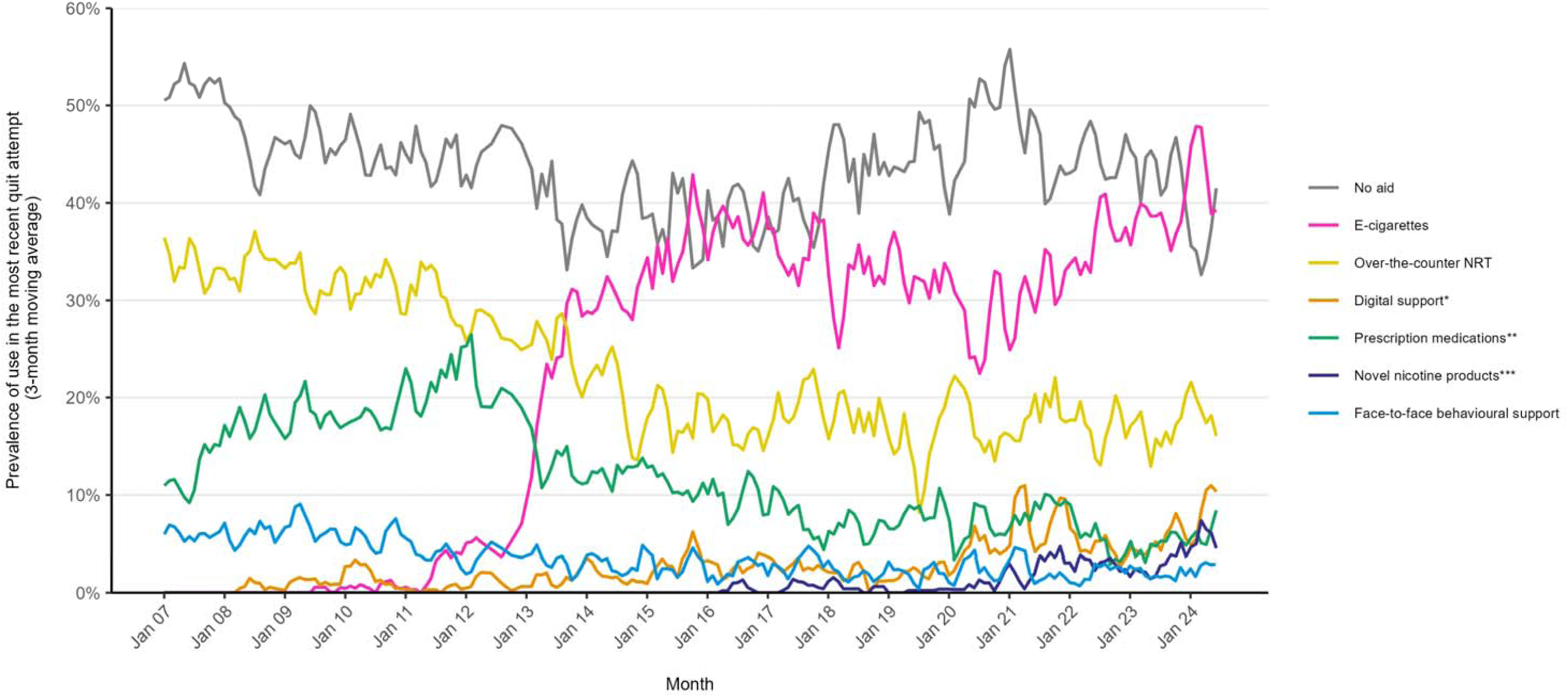
Monthly prevalence of the use of smoking cessation aids in quit attempts in England * Digital support includes websites and smartphone apps. ** Prescription medications include prescription nicotine replacement therapy, varenicline, and bupropion. *** Novel nicotine products include heated tobacco products and nicotine pouches. Corresponding figures showing data separately for each of these aids, and other aids not presented here (written self-help materials, telephone support, Allen Carr’s Easyway method, and hypnotherapy) are provided in **Figure S1.**

### Associations with abstinence

Overall, 17.7% [17.2-18.2%] of participants reported continuous abstinence from the start of their most recent quit attempt up to the time of the survey. **Table 2** shows unadjusted self-reported quit rates and sequentially adjusted models testing associations between each cessation aid and abstinence. Unadjusted quit rates were highest among users of nicotine pouches (30.1%), followed by heated tobacco products (29.9%), smartphone apps (28.0%), e-cigarettes (23.9%), and websites (23.4%).

**Table 2.** Associations of use of cessation aids with successful quitting.

| Cessation aid used<sup>1</sup> | Unadjusted quit rate, % [95% CI] | Odds of quit success, OR [95% CI] |  |  | Interaction with social grade<sup>5</sup> | Bayes factor<sup>6</sup> |
| --- | --- | --- | --- | --- | --- | --- |
|  |  | Model 1<sup>2</sup> | Model 2<sup>3</sup> | Model 3<sup>4</sup> |  |  |
| Non-prescription nicotine products |  |  |  |  |  |  |
| E-cigarettes | 23.9 [22.6–25.3] | 1.51 [1.38–1.65] | 1.90 [1.70–2.11] | 1.95 [1.74–2.17] | 0.97 [0.79–1.18] | - |
| Over-the-counter NRT | 13.5 [12.5–14.4] | 0.70 [0.63–0.76] | 0.96 [0.87–1.06] | 1.03 [0.93–1.15] | 0.92 [0.76–1.13] | 0.22 |
| Nicotine pouches | 30.1 [19.3–40.8] | 1.74 [1.03–2.95] | 1.45 [0.84–2.52] | 1.21 [0.70–2.07] | 0.55 [0.18–1.63] | 0.95 |
| Heated tobacco products | 29.9 [18.3–41.5] | 1.84 [1.06–3.23] | 2.35 [1.25–4.42] | 2.37 [1.24–4.51] | 0.34 [0.09–1.36] | - |
| Prescription medications |  |  |  |  |  |  |
| Prescription NRT | 15.0 [13.1–16.8] | 0.82 [0.71–0.95] | 1.33 [1.13–1.58] | 1.33 [1.12–1.58] | 0.85 [0.61–1.19] | - |
| Varenicline | 19.7 [17.2–22.2] | 1.16 [0.99–1.37] | 1.67 [1.39–2.00] | 1.80 [1.50–2.18] | 1.00 [0.70–1.44] | - |
| Bupropion | 11.0 [7.5–14.6] | 0.59 [0.41–0.86] | 1.14 [0.76–1.69] | 1.11 [0.73–1.69] | 1.23 [0.51–2.97] | 0.68 |
| Behavioural support |  |  |  |  |  |  |
| Websites | 23.4 [19.0–27.9] | 0.73 [0.54–0.98] | 1.62 [1.20–2.19] | 1.43 [1.03–1.98] | 1.68 [0.91–3.09] | - |
| Smartphone apps | 28.0 [21.0–34.9] | 1.53 [1.06–2.20] | 1.41 [0.92–2.16] | 1.10 [0.71–1.71] | 1.30 [0.56–2.98] | 0.67 |
| Written self-help materials | 14.0 [10.5–17.4] | 1.31 [1.00–1.70] | 0.76 [0.55–1.04] | 0.73 [0.53–1.00] | 0.93 [0.50–1.74] | 0.13 |
| Face-to-face behavioural support | 16.7 [14.2–19.3] | 1.04 [0.86–1.26] | 1.38 [1.11–1.71] | 1.26 [1.01–1.58] | 1.62 [1.05–2.50] | - |
| Social grades ABC1 | 15.9 [12.1–19.8] | 0.81 [0.60–1.09] | 1.05 [0.76–1.45] | 0.91 [0.65–1.29] | - | 0.28 |
| Social grades C2DE | 17.3 [13.8–20.7] | 1.26 [0.98–1.62] | 1.72 [1.30–2.27] | 1.59 [1.19–2.14] | - | - |
| Telephone support | 17.0 [11.4–22.6] | 0.99 [0.65–1.49] | 1.23 [0.79–1.93] | 0.93 [0.58–1.50] | 0.44 [0.17–1.14] | 0.42 |
| Allen Carr's Easyway | 13.1 [2.7–23.5] | 0.68 [0.27–1.73] | 0.82 [0.25–2.73] | 0.73 [0.20–2.70] | 0.64 [0.05–9.07] | 0.71 |
| Alternative treatments |  |  |  |  |  |  |
| Hypnotherapy | 14.5 [9.6–19.4] | 0.80 [0.53–1.20] | 0.81 [0.54–1.21] | 0.79 [0.52–1.22] | 1.91 [0.86–4.24] | 0.26 |
| None of these | 18.0 [17.3–18.8] | - | 0.67 [0.62–0.73] | - | - | - |

CI, confidence interval. NRT, nicotine replacement therapy. OR, odds ratio.
<sup>1</sup> Sorted within categories from the most to least popular in 2023/24.
<sup>2</sup> Adjusted for use of all other cessation aids, but no covariates.
<sup>3</sup> Adjusted for covariates, but no other cessation aids.
<sup>4</sup> Fully adjusted for all cessation aids plus covariates.
<sup>5</sup> Higher ORs indicate greater effectiveness (and lower ORs indicate lower effectiveness) of the smoking cessation aid among those from less advantaged social grades (C2DE) compared with those from more advantaged social grades (ABC1).
<sup>6</sup> Bayes factors for non-significant associations in model 3, based on an expected effect size of OR=1.5. Values ≤1/3 support the null hypothesis (i.e., no difference in abstinence between use and non-use of the aid) and values between 1/3 and 3 suggest the data are insensitive.

Analyses that adjusted for use of other cessation aids, but no covariates (Model 1, **Table 2**) indicated that smokers who used heated tobacco products, nicotine pouches, smartphone apps, e-cigarettes, and written self-help materials in their most recent quit attempt had higher odds of abstinence than those who did not use these cessation aids. Those who used bupropion, over-the-counter NRT, websites, and prescription NRT had lower odds. Odds were similar for users vs. non-users of varenicline, face-to-face behavioural support, telephone support, Allen Carr’s Easyway method, and hypnotherapy.

After adjustment for sociodemographic variables, level of addiction, factors relating to the quit attempt, month and year of the survey, and mode of data collection, but excluding adjustment for other cessation aids (Model 2, **Table 2**), the odds of abstinence were higher among those who used heated tobacco products, e-cigarettes, varenicline, websites, face-to-face behavioural support, or prescription NRT, and lower among those who tried to quit unaided. A similar pattern of results was observed when use of other cessation aids was adjusted for (Model 3, **Table 2**).

Figure 2 displays the fully adjusted results (Model 3), sorted by the odds ratio (OR) and by the lower 95% CI. In each case, the three aids associated with the highest odds of abstinence were e-cigarettes (OR=1.95 [1.74-2.17]), varenicline (OR=1.80 [1.50-2.18]), and heated tobacco products (OR=2.37 [1.24-4.51]). While heated tobacco products had the largest OR, it also had the widest 95% CI (on account of the small number of participants using this aid; *n*=72), meaning the size of the association was less certain. Positive associations were more modest for websites (OR=1.43 [1.03-1.98]), prescription NRT (OR=1.33 [1.12-1.58]), and face-to-face behavioural support (OR=1.26 [1.01-1.58]).

**Figure 2.**
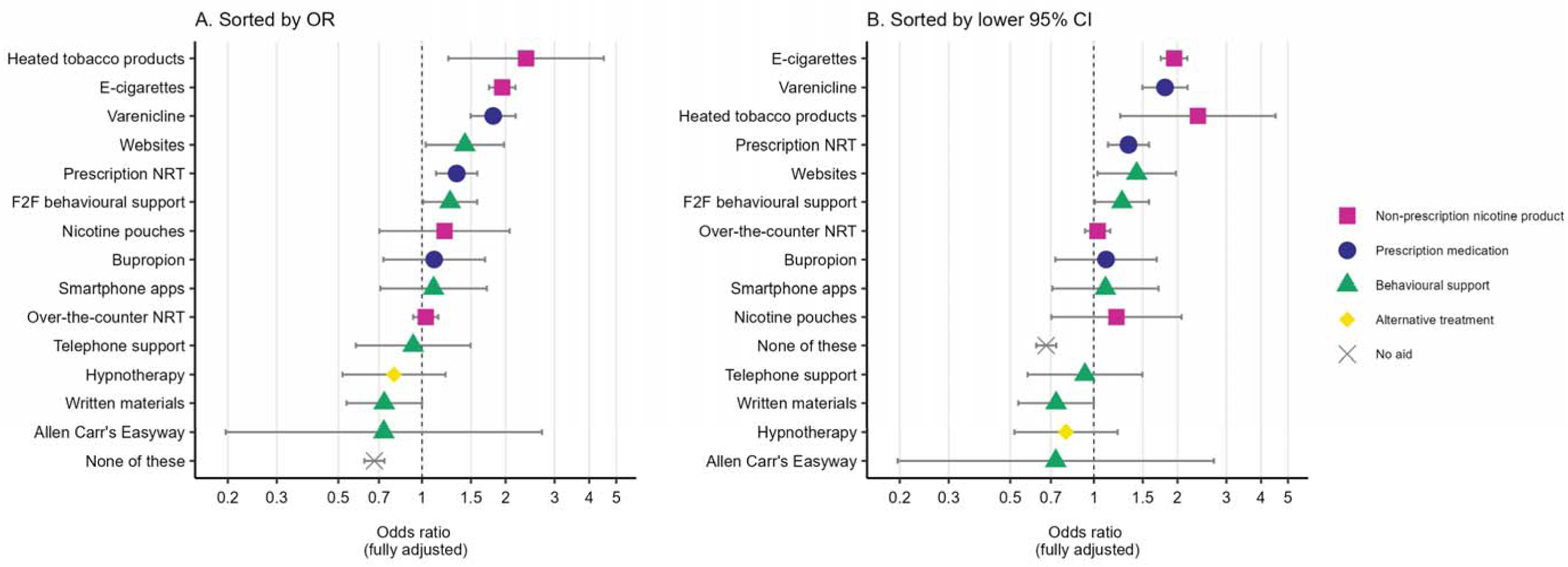
Fully adjusted associations between use of cessation aids and abstinence Data shown are the results of fully adjusted logistic regression models testing the association between use of a given aid and abstinence. Results for each individual aid are adjusted for use of all other cessation aids plus covariates (Model 3). Results for unaided quitting (i.e., ‘none of these’) are adjusted for covariates (Model 2).

There was little evidence that the effectiveness of the different cessation aids (fully adjusted for use of other aids and covariates) differed by social grade (**Table 2).** The only exception was use of face-to-face behavioural support, which was associated with higher odds of abstinence among those from less advantaged social grades (C2DE; fully adjusted OR=1.59 [1.19-2.14]) but similar odds of abstinence to non-use among those from more advantaged social grades (ABC1; fully adjusted OR=0.91 [0.65-1.29]; **Table 2**).

Bayes factors based on results from the fully adjusted model (**Table 2**) indicated that there was evidence to suggest there was no association between abstinence and use (vs. non-use) of over-the-counter NRT, face-to-face behavioural support for smokers from more advantaged social grades, written self-help materials, and hypnotherapy. The data were insensitive to detect associations between abstinence and use of nicotine pouches, bupropion, smartphone apps, telephone support, and Allen Carr’s Easyway method.

## Discussion

In England, many people who try to stop smoking do so without using any support. This is the least effective way to quit: our results show that after adjusting for their level of addiction and other features of their quit attempt, smokers who tried to quit unaided had around one-third lower odds of successfully quitting than those who used some form of support.

As of 2023/24, the most commonly used smoking cessation aid was e-cigarettes, used in 40% of quit attempts. After adjusting for covariates and use of other aids, we found that quit attempts aided by e-cigarettes were more likely to be successful than those that did not. This is consistent with evidence from randomised controlled trials^5,24^ and previous real-world studies^11,13^ and provides further evidence that, in addition to being popular, e-cigarettes offer one of the most effective methods of quitting smoking.

The next most popular aid in 2023/24 was over-the-counter NRT, used in 17% of quit attempts. While there is substantial trial evidence showing NRT to be an effective treatment^4^ (albeit less effective than e-cigarettes^5^), our data did not show any benefit of using over-the-counter NRT. We only found NRT to be associated with higher odds of success when it was obtained on prescription (which occurred much less frequently). This may be because when people buy NRT themselves, without any advice on how to use it effectively, they either do not use enough NRT or use it incorrectly.^25,26^ Evidence shows NRT is more effective for smoking cessation when used as a combination of a slow-release patch to suppress withdrawal symptoms and a fast-acting form (e.g., gum, lozenge, inhaler) to satisfy in-the-moment urges.^8,27^

Other aids positively associated with success in quitting were heated tobacco products, varenicline, websites, and face-to-face behavioural support, but these were used much less frequently (<5% of quit attempts in 2023/24). Of these, the effect estimate was largest for heated tobacco products, which were used in <1% of quit attempts in 2023/24, but the 95% CI was wide on account of the small number of participants who reported using them. It will be important to update this analysis when more data are available to improve the precision of our estimate.

Varenicline, which we found to be another of the most effective smoking cessation treatments, has not been available in England for several years. Its supply was paused in July 2021 after its manufacturer, Pfizer, detected higher than acceptable levels of nitrosamine impurities in the drug (a generic version of varenicline is available in some countries and is planned to be launched in late 2024 in the UK).^28^ The shortfall in varenicline use has not been offset by increases in the use of other prescription medications (NRT and bupropion).^28^ Even if it had, these medications appear to be less effective for helping people quit so they do not provide a like-for-like replacement.^8^ Cytisine, a similar compound to varenicline, started to be supplied on prescription via smoking cessation services from January 2024. Trial evidence suggests it is likely to be similarly effective to varenicline and e-cigarettes.^8^ We have been collecting data in the Smoking Toolkit Study on use of cytisine in quit attempts since April 2022 (prior to 2024, it was possible to buy it online or bring it into the UK from overseas) but there are not yet sufficient numbers of participants using cytisine to provide an estimate of its real-world effectiveness (*n*=9 as of June 2024). We aim to look at this in future when sample size allows.

Websites and smartphone apps have become more popular as quitting aids since 2020. This increase in the use of digital support coincided with the onset of the COVID-19 pandemic and may have been the result of people being less able to access in-person support,^29,30^ or less willing to go to pharmacies or shops to buy products such as NRT or e-cigarettes. In our previous analysis,^11^ which included data up to 2018, we found use of websites was associated with greater odds of quitting successfully among smokers from less (but not more) advantaged social grades. The present analysis showed a different pattern: increased odds of quitting among people who used websites compared with those who did not, with no evidence to suggest this association differed by social grade. This may potentially reflect changes in the profile of smokers using websites to quit in more recent years (as prevalence has increased), or improvements in the quality of information and support provided by the websites people are using.

This study is the first time we have examined the real-world effectiveness of smartphone apps. Our results did not show a clear benefit: while app use was associated with increased odds of success in partially adjusted models, the association was much smaller when we adjusted both for covariates and for use of other aids. Trial evidence on apps is also inconclusive.^31–33^ It is likely that effectiveness varies according to apps’ content^34^ and the extent to which they employ evidence-based behaviour change techniques.

Use of face-to-face behavioural support was associated with greater quit success among those from less advantaged social grades, but there was no association among those who were more advantaged. This was the only aid for which we observed a clear moderating effect of socioeconomic position. In England, this type of support is typically delivered by free-to-use, local authority-commissioned stop smoking services. These usually offer a combination of behavioural support and pharmacotherapy (e.g., varenicline, NRT, bupropion or in some services e-cigarettes); any benefits of the pharmacotherapy provided are accounted for in our models by adjustment for use of other aids. The difference in effectiveness by social grade may indicate that pharmacotherapy is sufficient for helping more advantaged smokers to stop smoking, but that for those who are less advantaged, face-to-face behavioural support adds value in terms of further increasing quit rates. Qualitative research with smokers of different socioeconomic positions who have used stop smoking services may be useful to gain insight into the elements people find most helpful.

There was not clear evidence of a benefit of any other aid, but some analyses were inconclusive. Although our overall sample size was large, some aids were used rarely across the entire period (bupropion, telephone support, and Allen Carr’s Easyway method) and some only started to become more popular late in the period (smartphone apps and nicotine pouches). As a result, there were only small numbers of users of these aids in our sample, which were not sufficient to draw firm conclusions. There is currently little evidence on the effectiveness of nicotine pouches,^35^ although their popularity appears to be rising quickly. Evidence on smartphone apps is also limited and findings are mixed (as outlined above).^31–33^ However, trials suggest bupropion, telephone support, and Allen Carr’s Easyway method increase quit rates under controlled trial conditions.^8,36–39^ Bayes factors indicated our data were insensitive to detect benefits of these five aids, meaning we need to collect more data to determine how effective they are in the real world. Our findings were clearer for written self-help materials and hypnotherapy: the data suggested using these aids was not associated with increased success in quitting. The lack of a clear benefit of written self-help materials contradicts trial evidence that shows that when no other support is available, using written self-help materials can increase quit rates.^40^ In our sample, around half of those who used written self-help materials (48.9%; 208/425) also used at least one other aid, which may account for some of the difference in results.

Strengths of this study include the representative sample, assessment of a wide range of smoking cessation aids, and adjustment for important confounders (e.g., level of addiction). There were also limitations. First, our outcome was based on self-reports of continuous abstinence, with no fixed duration required to determine success. As such, the definition of successful quitting varied across participants depending on how long ago their most recent past-year quit attempt started. This could potentially have caused us to overestimate success rates associated with aids that have become more popular very recently (e.g., nicotine pouches), if quit attempts involving these aids occurred closer to the time of the survey. Secondly, use of some aids was not assessed consistently across the entire period, so values were imputed with 0 in waves before data were collected. However, this is unlikely to have had a substantial impact on the results, given the aids that were introduced later in the period had very low prevalence before we started assessing them (e.g., e-cigarettes, heated tobacco products, nicotine pouches). Thirdly, as outlined above, although our overall sample size was large, analyses of some aids that were used more rarely were limited by small samples. This is therefore an analysis that will need to be updated again when more data are available. Finally, we only considered the effectiveness of aids for cessation and not other factors that may be important to consider when making decisions about which aid to use in a quit attempt (e.g., potential harms).

In conclusion, while a range of effective smoking cessation aids are available in England, many people try to quit either using less effective forms of support or none at all. Quit success rates could be improved by encouraging people to use more effective methods.

## Data Availability

All data produced in the present study are available upon reasonable request to the authors

## Declarations

### Ethics approval

Ethical approval was provided by the UCL Research Ethics Committee (0498/001). Participants provide informed consent to take part in the study, and all methods are carried out in accordance with relevant regulations. The data are not collected by UCL and are anonymised when received by UCL.

## References

1 West R. Tobacco smoking: Health impact, prevalence, correlates and interventions. Psychol Health 2017; 32: 1018–1036.

2 Hughes JR, Keely J, Naud S. Shape of the relapse curve and long-term abstinence among untreated smokers. Addiction 2004; 99: 29–38.

3 Hiscock R, Bauld L, Amos A, Fidler JA, Munafò M. Socioeconomic status and smoking: a review. Ann N Y Acad Sci 2012; 1248: 107–123.

4 Hartmann-Boyce J, Chepkin SC, Ye W, Bullen C, Lancaster T. Nicotine replacement therapy versus control for smoking cessation. Cochrane Database Syst Rev 2018. doi:10.1002/14651858.CD000146.pub5.

5 Lindson N, Butler AR, McRobbie H, Bullen C, Hajek P, Begh R et al. Electronic cigarettes for smoking cessation. Cochrane Database Syst Rev 2024. doi:10.1002/14651858.CD010216.pub8.

6 Hartmann-Boyce J, Livingstone-Banks J, Ordóñez-Mena JM, Fanshawe TR, Lindson N, Freeman SC et al. Behavioural interventions for smoking cessation: an overview and network meta-analysis. Cochrane Database Syst Rev 2021. doi:10.1002/14651858.CD013229.pub2.

7 Livingstone-Banks J, Fanshawe TR, Thomas KH, Theodoulou A, Hajizadeh A, Hartman L et al. Nicotine receptor partial agonists for smoking cessation. Cochrane Database Syst Rev 2023. doi:10.1002/14651858.CD006103.pub9.

8 Lindson N, Theodoulou A, Ordóñez-Mena JM, Fanshawe TR, Sutton AJ, Livingstone-Banks J et al. Pharmacological and electronic cigarette interventions for smoking cessation in adults: component network meta-analyses. Cochrane Database Syst Rev 2023. doi:10.1002/14651858.CD015226.pub2.

9 Munafò MR, Davey Smith G. Robust research needs many lines of evidence. Nature 2018; 553: 399–401.

10 Fidler JA, Shahab L, West O, Jarvis MJ, McEwen A, Stapleton JA et al. ‘The smoking toolkit study’: a national study of smoking and smoking cessation in England. BMC Public Health 2011; 11: 479.

11 Jackson SE, Kotz D, West R, Brown J. Moderators of real-world effectiveness of smoking cessation aids: a population study. Addiction 2019; 114: 1627–1638.

12 Jackson SE, Cox S, Shahab L, Brown J. Prevalence of use and real-world effectiveness of smoking cessation aids during the COVID-19 pandemic: a representative study of smokers in England. Addiction 2022; 117: 2504–2514.

13 Brown J, Beard E, Kotz D, Michie S, West R. Real-world effectiveness of e-cigarettes when used to aid smoking cessation: a cross-sectional population study. Addiction 2014; 109: 1531– 1540.

14 Kotz D, Brown J, West R. ‘Real-world’effectiveness of smoking cessation treatments: a population study. Addiction 2014; 109: 491–499.

15 Jackson SE, Kock L, Kotz D, Brown J. Real-world effectiveness of smoking cessation aids: A population survey in England with 12-month follow-up, 2015–2020. Addict Behav 2022; 135: 107442.

16 Kotz D, Brown J, West R. Prospective Cohort Study of the Effectiveness of Smoking Cessation Treatments Used in the “Real World”. Mayo Clin Proc 2014; 89: 1360–1367.

17 Kock L, Tattan-Birch H, Jackson S, Shahab L, Brown J. Socio-demographic, smoking and drinking characteristics in GB: A comparison of independent telephone and face-to-face Smoking and Alcohol Toolkit surveys conducted in March 2022. Qeios 2022. doi:10.32388/CLXK4D.

18 National Readership Survey. Social Grade. http://www.nrs.co.uk/nrs-print/lifestyle-and-classification-data/social-grade/ (accessed 3 Feb2019).

19 Fidler JA, Shahab L, West R. Strength of urges to smoke as a measure of severity of cigarette dependence: comparison with the Fagerström Test for Nicotine Dependence and its components. Addiction 2011; 106: 631–638.

20 Tattan-Birch H. BayesFactor.info. https://harry-tattan-birch.shinyapps.io/bayes-factor-calculator/ (accessed 28 May2024).

21 Dienes Z. Using Bayes to get the most out of non-significant results. Front Psychol 2014; 5: 781.

22 Jeffreys H. The theory of probability. OuP Oxford, 1998https://books.google.co.uk/books?hl=en&lr=&id=vh9Act9rtzQC&oi=fnd&pg=PA1&dq=The+theory+of+probability+jeffreys+1961&ots=fgQzMZX1lV&sig=q6SOaO0-cLW6gFlj8OOOUoRMZHc (accessed 28 May2024).

23 Lancaster T, Stead LF. Individual behavioural counselling for smoking cessation. Cochrane Database Syst Rev 2017. doi:10.1002/14651858.CD001292.pub3.

24 Auer R, Schoeni A, Humair J-P, Jacot-Sadowski I, Berlin I, Stuber MJ et al. Electronic Nicotine-Delivery Systems for Smoking Cessation. N Engl J Med 2024; 390: 601–610.

25 Beard E, Vangeli E, Michie S, West R. The Use of Nicotine Replacement Therapy for Smoking Reduction and Temporary Abstinence: An Interview Study. Nicotine Tob Res 2012; 14: 849– 856.

26 Mendelsohn C. Optimising nicotine replacement therapy in clinical practice. Aust Fam Physician 2013; 42: 305–309.

27 Theodoulou A, Chepkin SC, Ye W, Fanshawe TR, Bullen C, Hartmann-Boyce J et al. Different doses, durations and modes of delivery of nicotine replacement therapy for smoking cessation. Cochrane Database Syst Rev 2023. doi:10.1002/14651858.CD013308.pub2.

28 Jackson SE, Brown J, Tattan-Birch H, Shahab L. Impact of the disruption in supply of varenicline since 2021 on smoking cessation in England: A population study. Addiction 2024; 119: 1203–1210.

29 Action on Smoking and Health, Cancer Research UK. Stepping up: The response of stop smoking services in England to the Covid-19 pandemic. 2021https://ash.org.uk/wp-content/uploads/2021/01/ASH-CRUK-Stepping-Up-FINAL.pdf (accessed 5 May2021).

30 Healthwatch. GP access during COVID-19: A review of our evidence: April 2019 – December 2020. 2021https://www.healthwatch.co.uk/sites/healthwatch.co.uk/files/20210215%20GP%20access%20during%20COVID19%20report%20final_0.pdf (accessed 5 May2021).

31 Guo Y-Q, Chen Y, Dabbs AD, Wu Y. The Effectiveness of Smartphone App–Based Interventions for Assisting Smoking Cessation: Systematic Review and Meta-analysis. J Med Internet Res 2023; 25: e43242.

32 Whittaker R, McRobbie H, Bullen C, Rodgers A, Gu Y, Dobson R. Mobile phone text messaging and app-based interventions for smoking cessation. Cochrane Database Syst Rev 2019. doi:10.1002/14651858.CD006611.pub5.

33 Fang YE, Zhang Z, Wang R, Yang B, Chen C, Nisa C et al. Effectiveness of eHealth Smoking Cessation Interventions: Systematic Review and Meta-Analysis. J Med Internet Res 2023; 25: e45111.

34 Crane D, Ubhi HK, Brown J, West R. Relative effectiveness of a full versus reduced version of the ‘Smoke Free’ mobile application for smoking cessation: a randomised controlled trial. F1000Research 2018; **7**: 1524.

35 Patwardhan S, Fagerström K. The New Nicotine Pouch Category: A Tobacco Harm Reduction Tool? Nicotine Tob Res 2022; 24: 623–625.

36 Frings D, Albery IP, Moss AC, Brunger H, Burghelea M, White S et al. Comparison of Allen Carr’s Easyway programme with a specialist behavioural and pharmacological smoking cessation support service: a randomized controlled trial. Addiction 2020; 115: 977–985.

37 Keogan S, Li S, Clancy L. Allen Carr’s Easyway to Stop Smoking - A randomised clinical trial. Tob Control 2019; 28: 414–419.

38 Matkin W, Ordóñez Mena^a^ JM, Hartmann Boyce J. Telephone counselling for smoking cessation. Cochrane Database Syst Rev 2019.https://www.cochranelibrary.com/cdsr/doi/10.1002/14651858.CD002850.pub4/full (accessed 1 Aug2024).

39 Hajizadeh A, Howes S, Theodoulou A, Klemperer E, Hartmann-Boyce J, Livingstone-Banks J et al. Antidepressants for smoking cessation. Cochrane Database Syst Rev 2023.https://www.cochranelibrary.com/cdsr/doi/10.1002/14651858.CD000031.pub6/full?highlightAbstract=bupropion (accessed 1 Aug2024).

40 Livingstone-Banks J, Ordóñez-Mena JM, Hartmann-Boyce J. Print-based self-help interventions for smoking cessation. Cochrane Database Syst Rev 2019.https://www.cochranelibrary.com/cdsr/doi/10.1002/14651858.CD001118.pub4/full (accessed 20 Aug2024).

