## Supplementary material for "Prevalence and real-world effectiveness of popular smoking cessation aids in England: a population-based study": Table S1

### **Full details of measures**

#### Outcome: quit success

Past-year smokers who reported having made at least one serious quit attempt in the past year were asked ‘How long did your most recent quit attempt last before you went back to smoking?’ Responses were coded 1 for those who responded that they were ‘still not smoking’ and 0 otherwise.

#### Exposures: use of cessation aids

Past-year smokers who reported having made at least one serious quit attempt in the past year were asked: ‘Which, if any, of the following did you try to help you stop smoking
during the most recent serious quit attempt?’ Response options were coded as follows. Note that not all of these response options were included in every wave (some were introduced part-way through the study period [see Methods for details] and the wording of some response options changed slightly). For each aid, we coded anyone responding affirmatively to one or more of the relevant items as having used that aid (coded 1), else they were considered not to have used that aid (coded 0).

- Prescription NRT
  - ‘Nicotine replacement product on prescription or given to you by a health professional’
- NRT bought over the counter
  - ‘Nicotine replacement product (e.g. patches/gum/inhaler) without a prescription’
- Varenicline
  - ‘Champix (varenicline)’
- Bupropion
  - ‘Zyban (bupropion)’
- E-cigarettes
  - ‘Electronic cigarette'
  - ‘Electronic cigarette or vaping device’
  - ‘Juul’
- Face-to-face behavioural support
  - ‘Attended an NHS Stop Smoking group’
  - ‘Attended one or more NHS stop smoking one-to-one support sessions’
  - ‘Attended a non-NHS Stop Smoking group’
  - ‘Attended one or more non-NHS stop smoking one-to-one support sessions’
  - ‘Attended a Stop Smoking group’
  - Attended one or more Stop Smoking one-to-one counselling/advice/support ‘session/s’
- Telephone support
  - ‘Smoking helpline such as NHS smoking helpline or Quitline’
  - ‘Phoned NHS Smoking Helpline’
  - ‘Phoned a non-NHS smoking helpline’
  - ‘Phoned a smoking helpline’
- Written self-help materials
  - ‘Read a book/booklet’
  - ‘Allen Carr Easyway book’
  - ‘The SmokeFree Formula book’
  - ‘Other book or booklet’
- Websites
  - ‘A website’
  - ‘Visited www.nhs.uk/smokefree website’
  - ‘Visited a website other than Smokefree’
- Smartphone apps
  - ‘Used an application ('app') on a handheld computer (smartphone, tablet, PDA)’
- Hypnotherapy
  - ‘Hypnotherapy’
- Allen Carr’s Easyway method
  - ‘Allen Carr Easyway session’
- Heated tobacco products
  - ‘Heat-not-burn cigarette (e.g. iQOS with HEETS, heatsticks)’
- Nicotine pouches
  - ‘Tobacco-free nicotine pouch/pod or 'white pouches' that you place on your gum (e.g., Zyn, On!, Nordic Spirit, Velo, Lyft, Skruf)’

#### Covariates

Age was analysed as a categorical variable (16-24/25-34/35-44/45-54/55-64/≥65 years).

Gender was self-reported as man or woman. In more recent waves, participants have also had the option to describe their gender in another way; those who identified in another way were excluded due to low numbers.

Occupational social grade was categorised based on National Readership Survey classifications^1^ as ABC1 (includes managerial, professional, and upper supervisory occupations) and C2DE (includes manual routine, semi-routine, lower supervisory, state pension, and long-term unemployed). This occupational measure of social grade is a widely used and valid index of socioeconomic position that is widely used in research in UK populations. It has been identified as particularly relevant in the context of tobacco use and quitting.^2^

Level of addiction was assessed with two questions that asked participants to self-report ratings of the strength of urges to smoke over the last 24 hours.^3^ The first question asked: ‘How much of the time have you felt the urge to smoke in the past 24 hours?’ with response options:

1. Not at all
2. A little of the time
3. Some of the time
4. A lot of the time
5. Almost all of the time
6. All the time

Those who responded 2-6 were then asked: ‘In general, how strong have the urges to smoke been?’ with response options:

1. Slight
2. Moderate
3. Strong
4. Very strong
5. Extremely strong

We coded level of addiction as 0 for those who responded ‘not at all’ to the first question and as 1, 2, 3, 4, and 5 for those who responded ‘slight’, ‘moderate’, ‘strong’, ‘very strong’, and ‘extremely strong’ to the second question. This validated measure has similar predictive value as the Fagerström Test of Cigarette Dependence and the Heaviness of Smoking Index for cessation.^4^

Time since the quit attempt started was assessed with the question: ‘How long ago did your most recent serious quit attempt start? By most recent, we mean the last time you tried to quit’ with response options:

1. In the last week
2. More than a week and up to a month
3. More than 1 month and up to 2 months
4. More than 2 months and up to 3 months
5. More than 3 months and up to 6 months
6. More than 6 months and up to a year

We categorised responses as <1 month, 1-6 months, and >6 months.

The number of prior quit attempts in the past year was assessed with the question: ‘How many serious attempts to stop smoking have you made in the last 12 months? By serious attempt I mean you decided that you would try to make sure you never smoked again. Please include any attempt that you are currently making and please include any successful attempt made within the last year.’ We categorised responses as 1, 2, 3 or ≥4.

Whether or not the quit attempt was planned was assessed with the question: ‘Which one of the following applies to your most recent serious quit attempt?’ with response options:

1. I planned the quit for later the same day or for a date in the future
2. I started the quit attempt the moment I made the decision I was going to stop

Whether the participant cut down first or stopped abruptly was assessed with the question: ‘Did you cut down the amount you smoked before trying to stop completely at your most recent serious quit attempt?’ with response options:

1. Cut down first
2. Stopped without cutting down

The month and year of survey were included to account for seasonal variation in quit attempts and changes in the availability and regulation of different smoking cessation aids over the study period.

We also adjusted for the mode of data collection with a variable coded 0 up to February 2020 (when data were collected face to face) and 1 from April 2020 onwards (when data were collected via telephone).

**References**

1.	National Readership Survey. Social grade - definitions and discriminatory power. (2007).

2.	Kotz, D. & West, R. Explaining the social gradient in smoking cessation: it’s not in the trying, but in the succeeding. *Tob. Control* **18**, 43–46 (2009).

3.	West, R. J., Hajek, P. & Belcher, M. Severity of withdrawal symptoms as a predictor of outcome of an attempt to quit smoking. *Psychol. Med.* **19**, 981–985 (1989).

4.	Fidler, J. A., Shahab, L. & West, R. Strength of urges to smoke as a measure of severity of cigarette dependence: comparison with the Fagerström Test for Nicotine Dependence and its components. *Addict. Abingdon Engl.* **106**, 631–638 (2011).

### **Table S1.** Weighted sample characteristics

|  | Analysed sample (complete cases)^1^ | Excluded (missing data on ≥1 variable)^1^ |
| --- | --- | --- |
| Unweighted *N* | 25,094 | 1,695 |
| Age (years) |  |  |
| Mean (SD) | 38.7 (15.3) | 40.5 (16.2) |
| 16-24 | 21.5 [20.9–22.0] | 20.3 [18.2–22.5] |
| 25-34 | 25.3 [24.7–25.9] | 22.0 [19.9–24.3] |
| 35-44 | 20.2 [19.7–20.8] | 19.6 [17.6–21.8] |
| 45-54 | 15.6 [15.1–16.1] | 17.1 [15.2–19.2] |
| 55-64 | 10.5 [10.1–10.9] | 11.6 [10.1–13.4] |
| ≥65 | 7.0 [6.7–7.3] | 9.4 [8.0–10.9] |
| Missing^2^ | - | 3 |
| Gender |  |  |
| Men | 51.5 [50.8–52.2] | 49.4 [46.7–52.1] |
| Women | 48.5 [47.8–49.2] | 50.6 [47.9–53.3] |
| Missing^2,3^ | - | 93 |
| Occupational social grade |  |  |
| ABC1 (more advantaged) | 42.3 [41.7–43.0] | 40.1 [37.6–42.7] |
| C2DE (less advantaged) | 57.7 [57.0–58.3] | 59.9 [57.3–62.4] |
| Level of addiction^4^ |  |  |
| Not at all | 18.5 [18.0–19.0] | 23.3 [20.0–26.9] |
| Slight | 14.9 [14.4–15.4] | 14.7 [12.0–17.9] |
| Moderate | 41.2 [40.5–41.9] | 32.9 [29.1–36.9] |
| Strong | 17.7 [17.2–18.2] | 20.3 [17.1–23.9] |
| Very strong | 5.6 [5.3–5.9] | 6.3 [4.5–8.7] |
| Extremely strong | 2.1 [1.9–2.3] | 2.6 [1.6–4.1] |
| Missing^2^ | - | 1,045 |
| Time since the quit attempt started |  |  |
| <1 month | 15.8 [15.3–16.3] | 15.9 [14.0–18.1] |
| 1-6 months | 46.8 [46.1–47.5] | 49.2 [46.4–51.9] |
| >6 months | 37.4 [36.7–38.1] | 34.9 [32.3–37.5] |
| Missing^2^ | - | 152 |
| Number of past-year quit attempts |  |  |
| 1 | 65.0 [64.3–65.6] | 64.3 [61.8–66.8] |
| 2 | 21.0 [20.5–21.6] | 19.9 [17.9–22.0] |
| 3 | 7.6 [7.3–8.0] | 7.9 [6.6–9.5] |
| 4 or more | 6.4 [6.1–6.7] | 7.9 [6.6–9.5] |

Table continues on next page.

Table S1. continued

|  | Analysed sample (complete cases)^1^ | Excluded (missing data on ≥1 variable)^1^ |
| --- | --- | --- |
| Whether quit attempt was… |  |  |
| Planned | 46.0 [45.4–46.7] | 45.6 [42.7–48.6] |
| Unplanned | 54.0 [53.3–54.6] | 54.4 [51.4–57.3] |
| Missing^2^ | - | 390 |
| Gradual | 44.9 [44.3–45.6] | 45.1 [42.4–47.8] |
| Abrupt | 55.1 [54.4–55.7] | 54.9 [52.2–57.6] |
| Missing^2^ | - | 184 |
| Calendar month^5^, mean (SD) | 6.4 (3.5) | 6.3 (3.4) |
| Year^6^, mean (SD) | 2,014.0 (5.3) | 2,015.5 (4.9) |
| Telephone interview | 20.3 [19.7–20.8] | 34.0 [31.5–36.5] |

^1^ Data are presented as percentages with 95% confidence intervals, unless otherwise specified.

^2^ Unweighted number of missing cases.

^3^ Includes participants who described their gender ‘in another way’ (*n*=66).

^4^ Self-rated strength of urges to smoke over the past 24 hours.

^5^ Coded from January=1 to December=12.

^6^ Coded from 2006 to 2024.

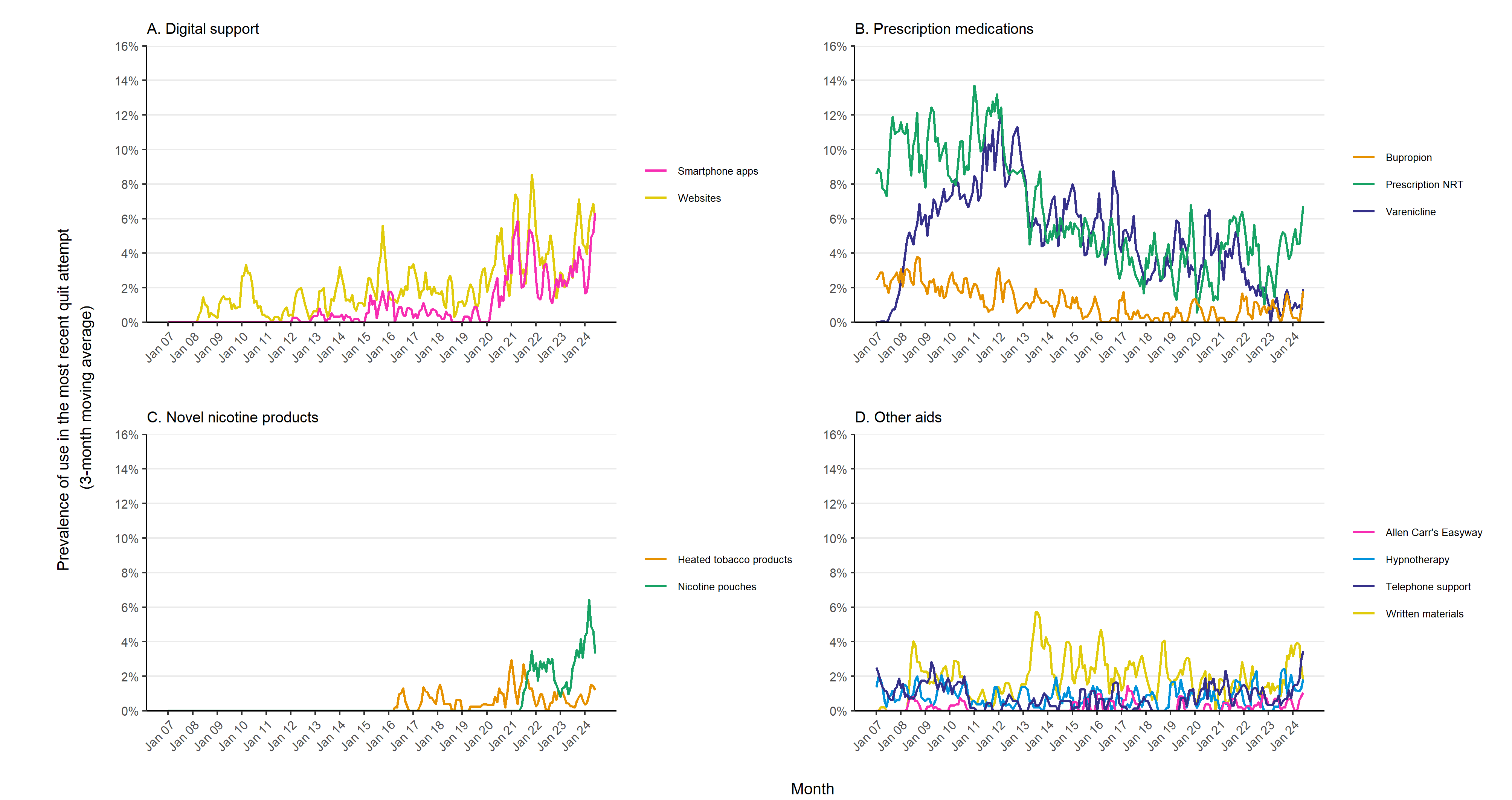

### **Figure S1. Monthly prevalence of the use of smoking cessation aids in quit attempts in England**

Corresponding data for other aids not presented here, and for digital support (websites and smartphone apps combined), prescription medications (nicotine replacement therapy, varenicline, and bupropion combined), and novel nicotine products (heated tobacco products and nicotine pouches combined) are provided in **Figure 1**.
